# Modern lineages of *Mycobacterium tuberculosis* were recently introduced in western India and demonstrate increased transmissibility

**DOI:** 10.1101/2022.01.04.22268645

**Authors:** Avika Dixit, Anju Kagal, Yasha Ektefaie, Luca Freschi, Rajesh Karyakarte, Rahul Lokhande, Matthias Groschel, Jeffrey A Tornheim, Nikhil Gupte, Neeta N Pradhan, Mandar S Paradkar, Sona Deshmukh, Dileep Kadam, Marco Schito, David M. Engelthaler, Amita Gupta, Jonathan Golub, Vidya Mave, Maha Farhat

**Author notes:** These authors contributed equally to this work (Vidya Mave, Maha Farhat).

## Abstract

**Background:** *Mycobacterium tuberculosis* (*Mtb*) transmissibility may vary between lineages (or variants) and this may contribute to the slow decline of tuberculosis (TB) incidence. The objective of our study was to compare transmissibility across four major lineages (L1-4) of *Mtb* among participants from two cohort studies in Pune, India.

**Methods:** We performed whole-genome sequencing (WGS) of *Mtb* sputum culture-positive isolates from participants in two prospective cohort studies of adults with pulmonary TB seeking care at public treatment centers in Pune, Maharashtra. We performed genotypic susceptibility prediction for both first- and second-line drugs using a previously validated random forest model. We used single nucleotide substitutions (SNS) and maximum likelihood estimation to build isolate phylogenies by lineage. We used Bayesian molecular dating to estimate ancestral node ages and compared tree characteristics using a two-sample Kolmogorov-Smirnov (KS) test.

**Results:** Of the 642 isolates from distinct study participants that underwent WGS, 612 met sequence quality criteria. The median age of the 612 participants was 31 years (IQR 24.4-44.2), the majority were male (64.7%) and sputum smear-positive (83.3%), and 6.7% had co-infection with HIV. Most isolates belonged to L3 (44.6%). The majority (61.1%) of multidrug-resistant isolates (MDR, resistant to isoniazid and rifampin) belonged to L2 (*P* < 0.001 [Fisher’s Exact]). There was no significant difference in host characteristics between participants infected with the four major lineages. In phylogenetic analysis, we measured shorter terminal branch lengths in the L2 tree compared to L1 and L3 trees indicating less time elapsing between transmission and sampling and higher transmissibility (median branch lengths: L2 - 3.3, L3 - 7.8, p <0.001). Branching times for L2 and L4 were more recent than L1 and L3 indicating recent introduction into the region (p < 0.01 [KS test]).

**Conclusion:** Modern *Mtb* lineages (L2 and L4) were more recently introduced in western India, compared to older lineages (L1 and L3). L2 shows a higher frequency of drug-resistance and higher transmissibility. Our findings highlight the need for contact tracing around cases of TB due to L2, and heightened surveillance of TB antibiotic resistance in India.

## Introduction

Tuberculosis (TB) has been the leading infectious disease killer globally, overtaken only recently by Coronavirus Disease 2019 (COVID-19)^1^. Despite investment in TB healthcare infrastructure, the decline in global incidence has been very slow (~2% annually)^1^, decelerated by ongoing community transmission^2,3^. In addition to host-related factors, *Mycobacterium tuberculosis* (*Mtb*) strain or lineage characteristics affect TB transmission^4^. An improved understanding of these characteristics is necessary to improve our ability to control TB transmission, especially in high burden settings.

The *Mtb* species, in its strictest sense (*sensu stricto*), has been divided into four major lineages based on genotype, the ancient lineage 1 (L1) and modern L2, 3, and 4^5^. Previous studies have supported transmissibility differences between these lineages based on the genetic similarity between transmitted isolates, increasing frequency of a lineage in a community, or lineage propensity for younger hosts (reflecting recent transmission)^6^. Attempts to characterize transmissibility differences between lineages has thus far been largely limited to the use of traditional molecular methods such as spoligotyping that underestimate genetic variation, may misclassify lineage assignment^7^ and overestimate transmission links^4^. Even with the use of modern methods, i.e. whole-genome sequencing (WGS) that can overcome the majority of these shortcomings, studies yielded conflicting results^8–11^. This is because they either lacked epidemiological data to control for exposure^8–10^, or were analyzing outbreak events caused by a single lineage and thus were limited by a lack of lineage diversity^11^.

India carries a quarter of the global burden and the highest annual incidence of TB^1^. TB in India is a microcosm of the global disease epidemic with increasing rates of antibiotic resistance in certain districts^12,13^ and continued community transmission of TB^2,3^. Although geographic variation in lineage prevalence has been noted, L1 comprises approximately two thirds (67%) of the *Mtb* isolates in the country, but all four major lineages are found in circulation^14^. Here, we leverage two prospective cohort studies of pulmonary TB in western India to assess transmission differences between the four major *Mtb* lineages using WGS between 2013 – 2018: (1) a prospective observational study of active TB in Pune, India^15^, and (2) a prospective study to estimate co-prevalence of diabetes and TB and its impact on TB treatment outcomes in Pune^16,17^. We study the frequency of drug resistance among the major *Mtb* lineages and explore the age of resistance acquisition, as well as discrepancies between genotype and phenotypic resistance discrepancy in this less studied TB patient population.

## Methods

### Study Design and Participant Eligibility

The WGS used in this study were obtained from: 1) the prospective Cohort for Tuberculosis Research by the Indo-US Medical Partnership (CTRIUMPH) study conducted in Pune, India from August 2014 to December 2019^15^, and 2) a prospective study, focused on the relationship between tuberculosis and diabetes in adults, conducted in Pune, India between December 2013 and May 2019^16,17^. The eligibility criteria and study procedures have been previously described^15–18^. Briefly, both studies enrolled participants with new active pulmonary TB (PTB) or extrapulmonary TB (EPTB) presenting to eleven TB treatment centers (run through the National Tuberculosis Elimination Program [NTEP], previously known as Revised National TB Control Programme [RNTCP]) located in the western Indian city of Pune, India. Any adult (>=18 years of age) participant with microbiologically confirmed pulmonary TB was included in this analysis. CTRIUMPH also enrolled household contacts (including those <18 years of age) of index pulmonary TB cases and followed them for 24 months for development of TB infection or TB disease. Microbiological confirmation could be through a positive result on any of the following: sputum smear for acid-fast bacilli (AFB), GeneXpert MTB/RIF (Cepheid, Sunnyvale, CA, USA), AFB culture, meeting criteria for clinical TB using NTEP guidelines. Information was collected at enrollment including, sociodemographic characteristics, past medical history including history of TB, diabetes mellitus, co-infection with human immunodeficiency virus (HIV). All study participants underwent clinical evaluation including anthropometrics, laboratory testing, and chest radiography.

### Culture and Phenotypic Drug Susceptibility Testing

Sputum samples were subjected to culture in Mycobacterial Growth Indicator Tube (MGIT) liquid culture and Lowenstein Jensen (LJ) media. Phenotypic drug susceptibility testing (DST) was performed for isoniazid, rifampin, ethambutol, and pyrazinamide using MGIT 960. We repeated phenotypic DST for isolates where the initial testing did not have concordance with genotypic DST. The critical concentrations used for MGIT 960 were: Isoniazid - 0.10ug/mL, Rifampin - 1.00ug/mL, Ethambutol - 5.00ug/mL, and Streptomycin - 1.00ug/mL.

### Whole-genome sequencing and variant identification

Positive LJ cultures from baseline sputum samples were subjected to deoxyribonucleic acid (DNA) extraction using a standardized protocol. Briefly, DNA was extracted using Ultra-Deep Microbiome Prep kit (Molzym, Bremen, Germany) version 2.1 with the following steps: 1) removal of other DNA, 2) lysis of pathogen, 3) DNA purification 4) DNA elution. Extracted DNA was collected in elution tubes and preserved at minus 200 °C for shipment and sequencing. Approximately one microgram of DNA per sample was fragmented using a Q800R2 sonicator (QSonica, Newtown, CT, USA) with the following parameters: 3 minutes sonication with 15 seconds pulse on, 15 seconds pulse off, and 20% amplitude. The fragmented DNA was size selected to target 600-650bp by fragment separation using the Agencourt AMPure XP beads (Beckman Coulter, Code A63882). DNA Library preparations for WGS were performed using the NEBNext® Ultra™ II DNA Library Prep Kit for Illumina® (New England BioLabs, Code E7645L) with the following modifications. The adapters and 8bp index oligos purchased from IDT® (Integrated DNA Technologies, San Diego, CA) based on Kozarewa and Turner, 2011^19^, were used in place of those supplied in the NEB preparation kit. A dual-indexing approach was also utilized^20^. The samples were sequenced on an Illumina NextSeq using a 300 cycle v2 mid output kit (Illumina, Code FC-404-2003) with the standard Illumina® procedure. The appropriate sequencing primers were added to the cartridge as in Kozarewa and Turner, 2011^19^. The raw reads were processed through a custom bioinformatics pipeline^21^. This pipeline uses PRINSEQ with an average Phred score threshold of 20 to trim the reads^22^, Kraken^23^ to confirm that reads belong to the *Mtb* complex excluding any isolate with <90% mapping. Reads are then aligned to the reference genome H37Rv (GenBank NC000962.3) using BWA MEM^24^, duplicate reads are removed using PICARD^25^, and variants are called using Pilon^26^. Isolate lineage was called using standard techniques^27–29^ implemented in a bioinformatics pipeline^30^.

### Genotypic drug susceptibility determination

We subjected the WGS for each isolate to genotypic drug susceptibility prediction using a random forest classifier^31^. This classifier predicts resistance to 13 anti-tubercular antibiotics using 992 known resistance-associated genetic variants and was recently validated against other genotypic resistance predictors and was found to have similar performance with a sensitivity of >90% for isoniazid and rifampin^32^.

### Phylogenetic analysis and comparison across lineages

A multiple sequence alignment was generated for isolates belonging to lineages 1-4. Isolates not belonging to these *Mtb sensu stricto* lineages were excluded. If >95% of the sequenced isolates did not have coverage of at least 10x at a site, the site was excluded from the alignment. We excluded variants in genes implicated in drug resistance from the phylogenetic construction,^33^ because drug resistance genes are under selective pressure and may bias tree structure. We also excluded insertions and deletions (‘indels’), transposases, and genes coding for the amino acid patterns proline-glutamate (PE) or proline-proline-glutamate (PPE)^34^ by convention. We used RAxML 8.2.11^35^ to generate a maximum likelihood (ML) tree, using H37Rv (GenBank NC000962.3) as the outgroup with each tree bootstrapped 1000 times. We assumed a general time-reversible nucleotide substitution model with the Γ distribution to model site rate heterogeneity^36^. We used BEAST v1.10.4^37^ to date the ML tree, assuming a relaxed molecular clock with a log-normal distribution and a mean rate of 0.5 SNS per genome per year based on prior published data^38^. We combined the outputs of the BEAST and bootstrap analyses using Sumtrees.py from the DendroPy version 4.5.2 library^39^. This resulted in a final dated phylogenetic tree with nodal bootstrap support that was visualized and annotated using Interactive Tree of Life^40^. Subsequent analysis was performed in R version 3.6.1 using *ape*^41^ and *phangorn*^42^ packages. Participant characteristics were compared across the four lineages using analysis of variance (ANOVA) for continuous variables or χ^2^ test for categorical variables. We compared the following characteristics across the four lineages using a two-sample Kolmogorov-Smirnov (KS) test: branch lengths, pairwise single nucleotide substitution (SNS) distances, and branching times, with a one-sided p-value of < 0.1 considered to be significant. We investigated possible transmission clusters using a range of cut-offs for SNS distance. For the clustering analysis, we defined clusters as isolates that had a maximum genetic distance using specified SNS thresholds. We used the *finalfit* R package version 1.03^43^ to perform logistic regression to identify factors associated with clustering at a SNS threshold of ≤25.

### Host contribution to transmission

We used a previously described ‘propensity to propagate’ (PTP) method^44^ to assess host-related transmission factors. Data were available on participant’s age, gender, smear positivity and alcohol use. All participants had pulmonary disease. Data were not available on drug use, occupation, and homelessness, while foreign birth did not apply to our setting. Alcohol use data were missing for two participants, and they were assumed to be non-consumers as the overall frequency of alcohol consumption was low.

## Results

### Study participant and Mtb isolate characteristics

Of the 2,257 participants screened in the two studies by 1^st^ July 2018, 1,046 (46%) had microbiologically positive sputum cultures at baseline. A total of 48 pediatric participants and 9 with EPTB who had positive cultures were excluded. Of these, a total of 880 (84%) study participants with *Mtb* growth on sputum culture at baseline were successfully sub-cultured. Only nine participants with active TB were identified among household contacts of index cases in the C-TRIUMPH study but were excluded from sequencing due to few numbers. Extracted DNA from index samples available as of February 1, 2019, (n=750) underwent WGS. Of these, 108 samples (14%) did not meet coverage threshold despite two attempts. Of the remaining 642 (86%), 612 (95%) met sequence quality criteria (**Methods**). The majority of the 612 sequenced participants were male (64.7%), and the median age of participants was 31 years (IQR: 24.4-44.2 years). Most participants had a positive sputum smear microscopy (83.3%); HIV co-infection was uncommon (6.7%); 13.1% had diabetes mellitus (DM). Participants were infected with L3 isolates most commonly (n=273, 44.7%), followed by L1 (n=162, 26.5%), L4 (n= 132, 21.6%), and L2 (n=45, 7.3%). Demographic characteristics and co-morbidities of study participants did not differ by *Mtb* lineage (**Table 1**). Among the 574 isolates that underwent phenotypic drug susceptibility testing, 49 (8.5%) were mono-resistant to isoniazid, 6 (1.0%) were mono-resistant to rifampin, and 24 (4.2%) were multi-drug resistant (MDR), i.e., resistant to both isoniazid and rifampin (**Supplementary Table 1**).

**Table 1:**
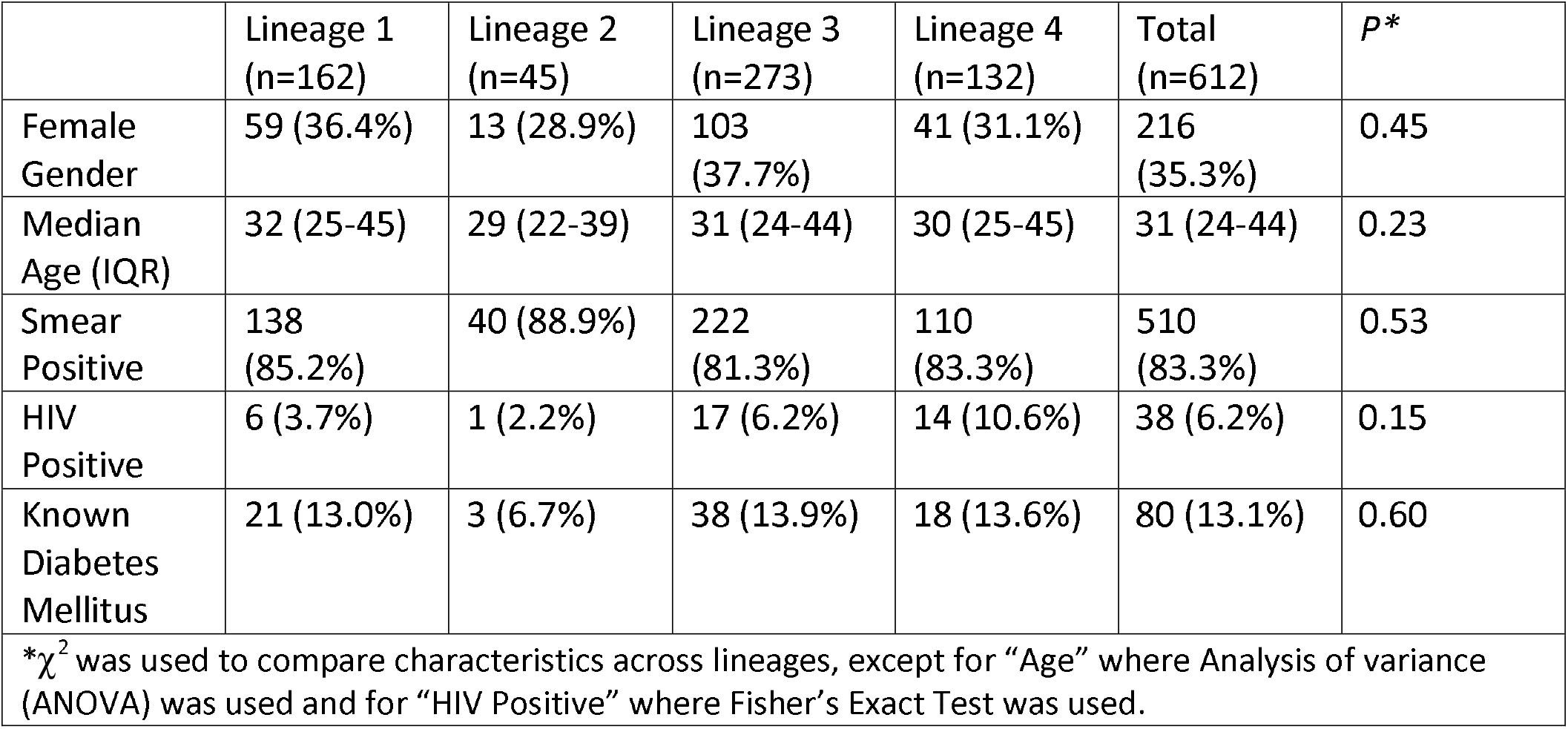
Demographic characteristics of study participants, by lineage.

|  | Lineage 1<br>(n=162) | Lineage 2<br>(n=45) | Lineage 3<br>(n=273) | Lineage 4<br>(n=132) | Total<br>(n=612) | *P* * |
| --- | --- | --- | --- | --- | --- | --- |
| Female Gender | 59 (36.4%) | 13 (28.9%) | 103 (37.7%) | 41 (31.1%) | 216 (35.3%) | 0.45 |
| Median Age (IQR) | 32 (25-45) | 29 (22-39) | 31 (24-44) | 30 (25-45) | 31 (24-44) | 0.23 |
| Smear Positive | 138 (85.2%) | 40 (88.9%) | 222 (81.3%) | 110 (83.3%) | 510 (83.3%) | 0.53 |
| HIV Positive | 6 (3.7%) | 1 (2.2%) | 17 (6.2%) | 14 (10.6%) | 38 (6.2%) | 0.15 |
| Known Diabetes Mellitus | 21 (13.0%) | 3 (6.7%) | 38 (13.9%) | 18 (13.6%) | 80 (13.1%) | 0.60 |

\* $\chi^2$ was used to compare characteristics across lineages, except for “Age” where Analysis of variance (ANOVA) was used and for “HIV Positive” where Fisher’s Exact Test was used.

### Genotypic drug susceptibility testing and concordance with phenotypes

We subjected the 612 *Mtb* isolates to genotypic drug susceptibility testing (DST) using a validated random forest model to assess discrepancies as these are understudied in the Indian context^31,32^. Fifteen isolates were concordant for rifampin resistance by phenotypic and genotypic DST; five tested resistant only by genotype, and 15 tested resistant only by phenotype. For isoniazid, 38 isolates were concordant for resistance; 35 were tested resistant only by phenotype, and 19 tested resistant only by genotype. We identified 24 (4.5%, n=533) isolates to be MDR based on phenotype and 18 (2.9%, n=612) isolates as MDR based on genotype (**Table 2**), of which 13 (2.1%, n=612) were concordant resistant by both methods.

**Table 2:** Isolates predicted to be resistant based on genotype, by drug and lineage.

|  | Isoniazid | Rifampin | Ethambutol | Ethionamide | Pyrazinamide | Streptomycin | Levofloxacin | Capreomycin | Multidrug resistant |
| --- | --- | --- | --- | --- | --- | --- | --- | --- | --- |
| **L1**<br>**(n=162)** | 11<br>(6.8) | 2 (1.2) | 0 | 5 (3.0) | 0 | 4 (2.5) | 2 (1.2) | 0 | 1 (0.6) |
| **L2**<br>**(n=45)** | 14<br>(31.1) | 11<br>(24.4) | 12<br>(26.7) | 4 (8.9) | 2 (4.4) | 17<br>(37.8) | 12<br>(26.7) | 0 | 11<br>(24.4) |
| **L3**<br>**(n=273)** | 23<br>(8.4) | 5 (1.8) | 5 (1.8) | 5 (1.8) | 4 (1.5) | 13<br>(4.8) | 10<br>(3.7) | 4 (1.5) | 4 (1.5) |
| **L4**<br>**(n=132)** | 12<br>(9.1) | 2 (1.5) | 0 | 2 (1.5) | 2 (1.5) | 8 (6.0) | 10<br>(7.6) | 0 | 2 (1.5) |
| **Total**<br>**(n=612)** | 60<br>(9.8) | 20<br>(3.3) | 17<br>(2.8) | 16<br>(2.6) | 8 (1.3) | 42<br>(6.9) | 34<br>(5.6) | 4 (0.7) | 18<br>(2.9) |
| **P**<br>**(Fisher's**<br>**Exact,**<br>**L2 vs**<br>**non-L2)** | <0.001 | <0.001 | <0.001 | 0.020 | 0.110 | <0.001 | <0.001 | - | <0.001 |

We investigated the discrepancies between genotypic and phenotypic DST since these are not well studied in India. We performed repeat DST for all discrepant isolates (n=98) for the drugs isoniazid (n=54), rifampin (n=20), ethambutol (n=26), and streptomycin (n=46). Of the 98, 64 (65.3%) isolates had growth on sub-cultures without contamination. Repeat phenotypic testing resolved a substantial number of discrepancies with few remaining discrepant isolates, isoniazid (n=6), rifampin (n=1), ethambutol (n=4), and streptomycin (n=8). Mutations found in these isolates are shown in **Supplementary Tables 2 and 3**. The 3 isolates identified as isoniazid resistant only by genotypic DST harbored canonical drug resistance-conferring mutation *katG* S315T. The one rifampin resistant isolate identified only on genotypic DST also harbored the canonical resistance-conferring mutation *rpoB* L430P. Mutations identified in isolates that were genotypically predicted resistant to ethambutol and streptomycin but were determined to be susceptible on phenotypic testing are shown in **Supplementary Table 2**. We conducted capreomycin phenotypic testing on four isolates that were predicted capreomycin resistant but susceptible to isoniazid and rifampicin based on genotype. All four of these isolates harbored a H68R mutation in the *tlyA* gene and tested phenotypically capreomycin susceptible at the critical concentration of 2.5 mcg/mL. Among genotypically susceptible and phenotypically resistant isolates (**Supplementary Table 3**), we found no mutations known to be associated with resistance. The isolates harbored 7 mutations not previously reported in the WHO resistance catalogue, and 13 mutations not associated with resistance or of unknown significance^45^.

### Phylogenetic characteristics of lineages

We generated a phylogenetic tree (**Figure 1**), and compared diversity and tree characteristics including terminal branch lengths as indirect measures of bacterial transmissibility as previously validated^9,29,46^ (**Methods**). The L2 phylogeny had shorter terminal branch lengths (median 3.3 [IQR: 1.3-9.2])) as compared to L1 & L3 (L1: 9.4 [5.4-15.7], p <0.001; L3: 7.8 [4.4-11.3], p <0.001) but not L4 (5.1 [0.8-10.5], p = 0.24) (**Figure 2A**). Isolates belonging to L2 were genetically more similar than those belonging to other lineages, as indicated by the smaller median pairwise SNS distance (L2: median 234, IQR: 215-279.5 *vs*. L1: median 865 SNSs, IQR: 496-944, p <0.001; L3: median 342, IQR: 306-372, p <0.001; L4: median 729, IQR: 651-771, p <0.001) (**Figure 2B**). Branching times for L2 (median 6.3 years, IQR: 3.2-10.9) were more recent than for L1 (median 12 years, IQR: 7-21, p < 0.001), L3 (median 10.4 years, IQR: 6.8-15.0, p = 0.001) but not L4 (median 9.0 years, IQR: 2.9-13.8, p = 0.071) (**Figure 2C**).

**Figure 1:**
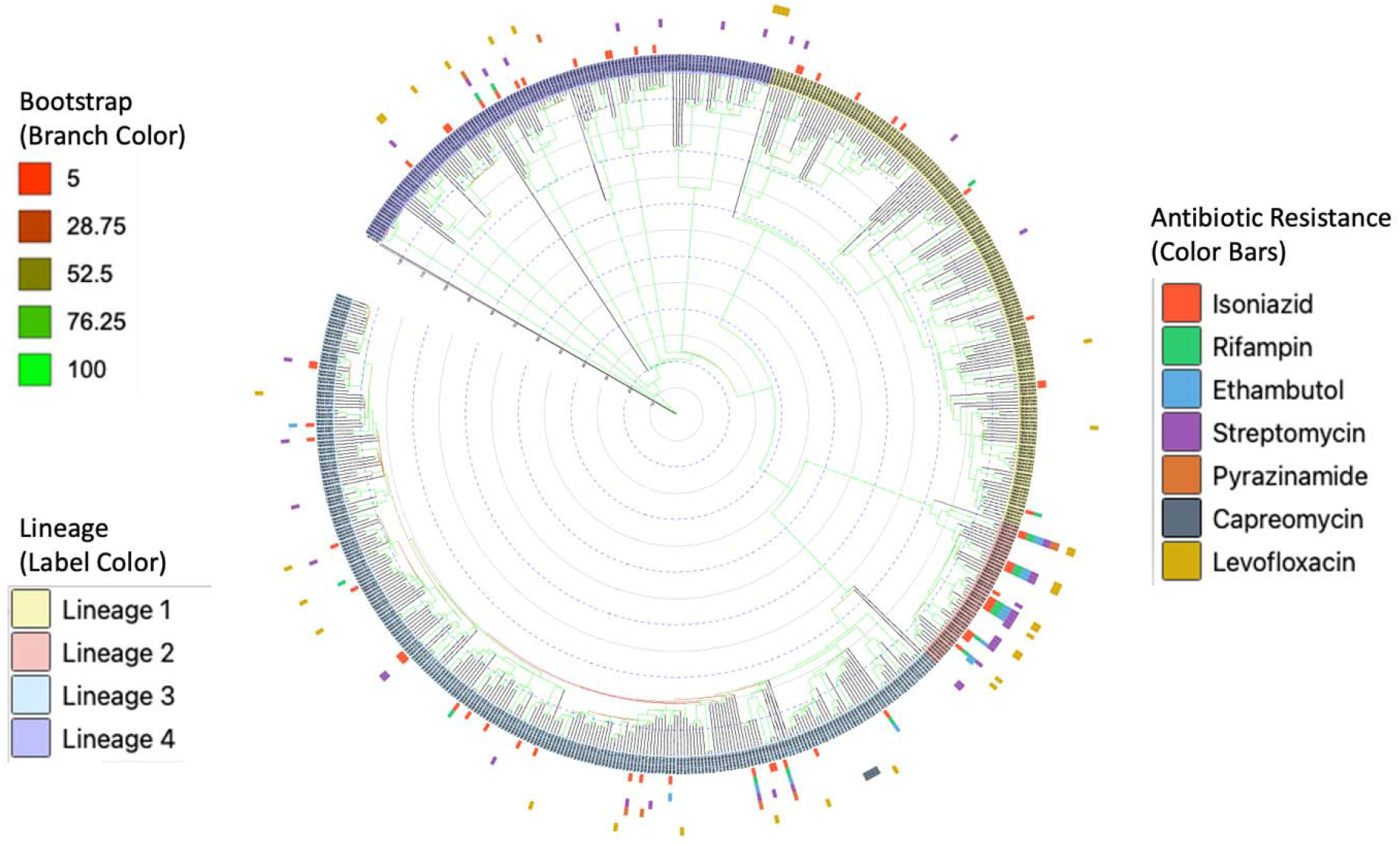
Dated Bayesian phylogenetic tree of 612 *Mycobacterium tuberculosis* isolates. Color bars represent antibiotic resistance and branch color represent bootstrap support. Node labels are shaded to depict lineage.

**Figure 2:**
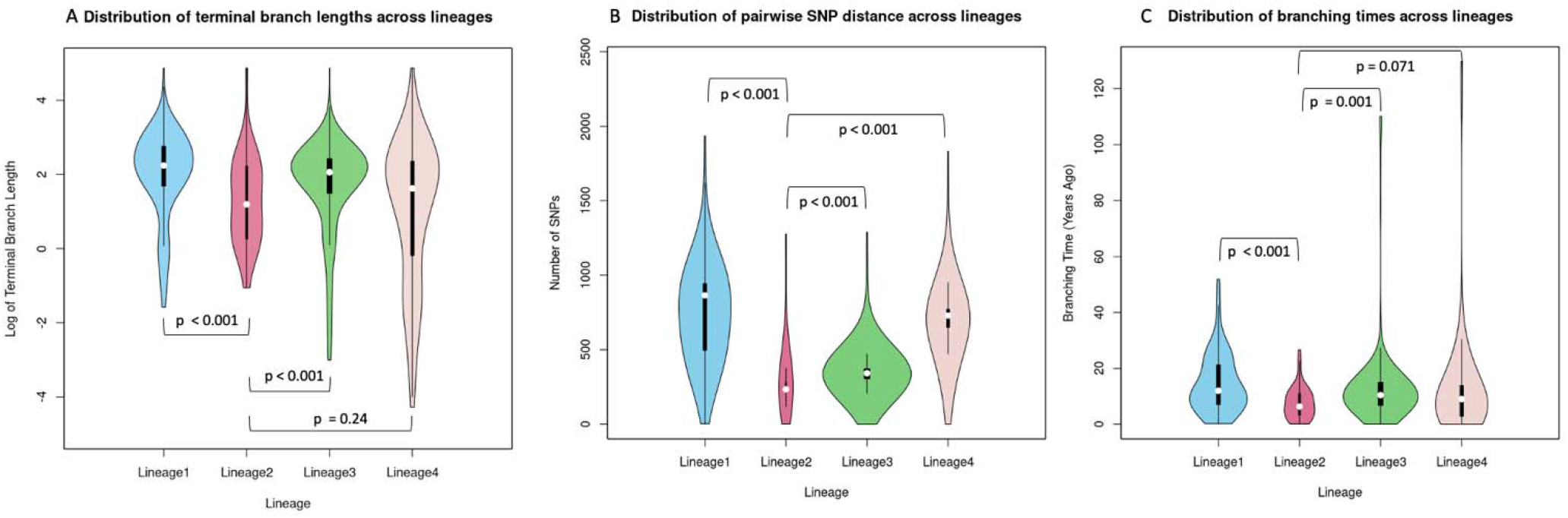
Lineage-wise distribution of A) terminal branch lengths, B) pairwise single nucleotide substitution (SNS) distance, and C) branching times, using 612 tuberculosis isolates from Pune, India. P values calculated using one-sided two-sample Kolmogorov-Smirnov test.

### Mtb clusters

We investigated possible transmission clusters using SNS cut-offs previously defined in the literature to investigate *Mtb* outbreaks^38^. We report results using the inclusive cut-off of less than or equal to 25 SNSs^38^ below, but the results are similar at lower thresholds and detailed in **Supplementary Table 4**. When evaluated cumulatively over a range of SNS distances spanning 1-100 SNSs, we found L2 & L4 to have a higher proportion of isolates belonging to clusters at any given SNS cut-off (**Figure 3**). We calculated the odds of an isolate being in a cluster (as defined by a threshold of ≤ 25 SNSs) and found that, as compared to L1, isolates belonging to L2 and L4 had more than 2- and 3-times the odds, respectively, of being in a cluster. The finding for L4 held even after multivariable control for host factors (**Table 3**).

**Figure 3:**
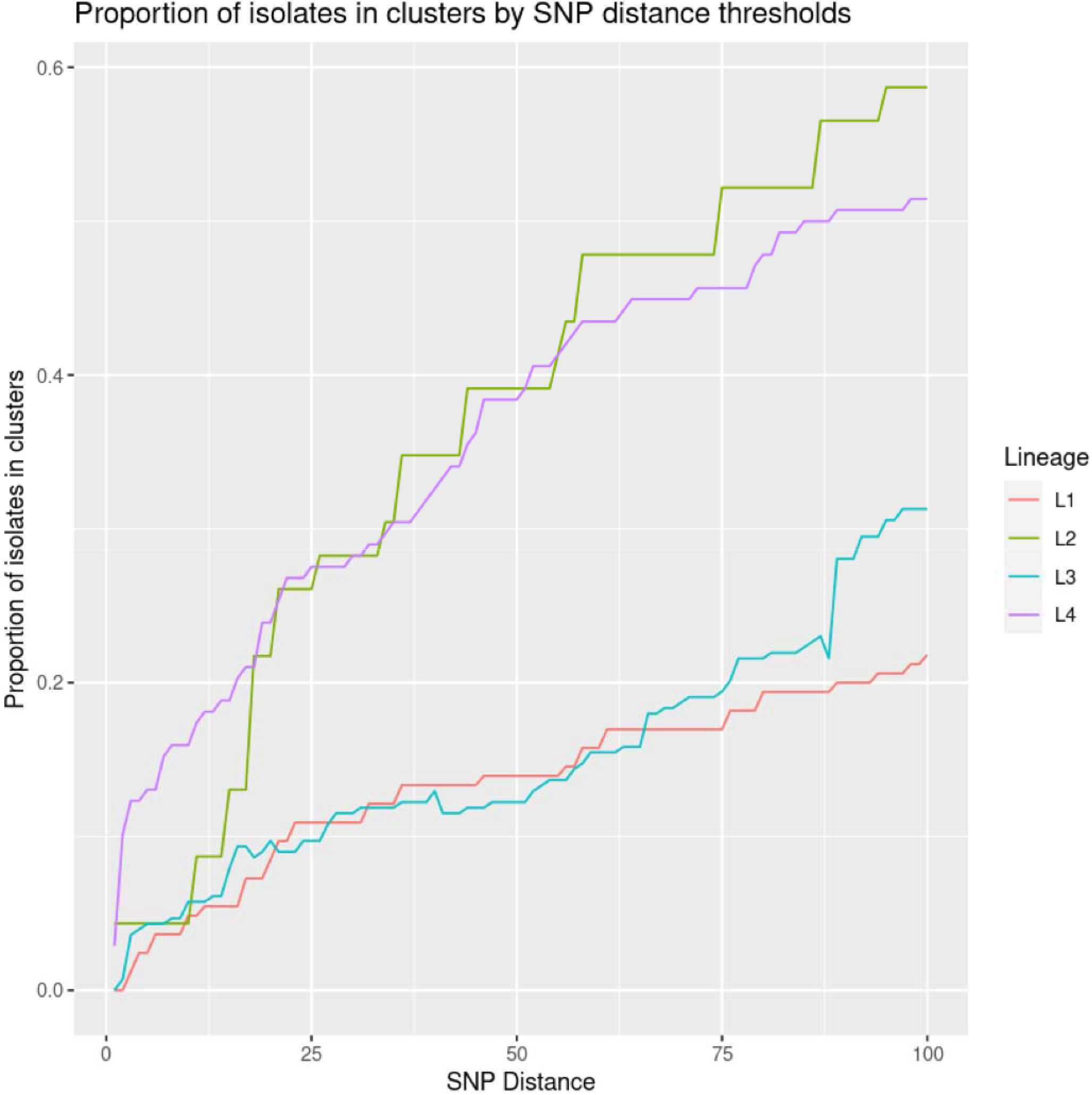
Proportion of isolates belonging to clusters by lineage (L) based on a range of single nucleotide substitution (SNS) distance threshold.

**Table 3:**
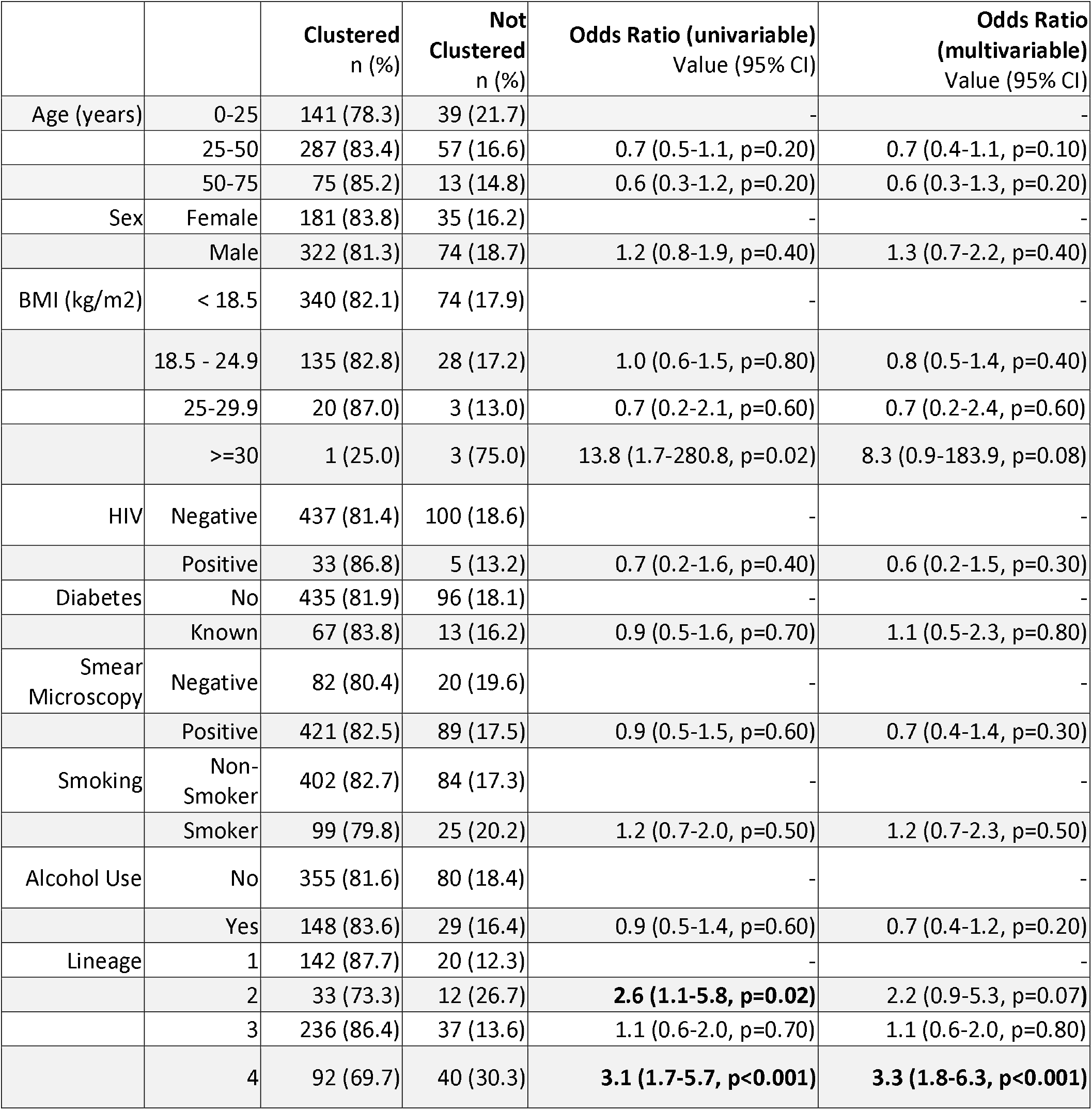
Characteristics associated with clustering. Clustering was defined as a single nucleotide substitution difference of less than or equal to 25.

|  |  | Clustered<br>n (%) | Not<br>Clustered<br>n (%) | Odds Ratio (univariable)<br>Value (95% CI) | Odds Ratio<br>(multivariable)<br>Value (95% CI) |
| --- | --- | --- | --- | --- | --- |
| Age (years) | 0-25 | 141 (78.3) | 39 (21.7) | - | - |
|  | 25-50 | 287 (83.4) | 57 (16.6) | 0.7 (0.5-1.1, p=0.20) | 0.7 (0.4-1.1, p=0.10) |
|  | 50-75 | 75 (85.2) | 13 (14.8) | 0.6 (0.3-1.2, p=0.20) | 0.6 (0.3-1.3, p=0.20) |
| Sex | Female | 181 (83.8) | 35 (16.2) | - | - |
|  | Male | 322 (81.3) | 74 (18.7) | 1.2 (0.8-1.9, p=0.40) | 1.3 (0.7-2.2, p=0.40) |
| BMI (kg/m<sup>2</sup>) | < 18.5 | 340 (82.1) | 74 (17.9) | - | - |
|  | 18.5 - 24.9 | 135 (82.8) | 28 (17.2) | 1.0 (0.6-1.5, p=0.80) | 0.8 (0.5-1.4, p=0.40) |
|  | 25-29.9 | 20 (87.0) | 3 (13.0) | 0.7 (0.2-2.1, p=0.60) | 0.7 (0.2-2.4, p=0.60) |
|  | >=30 | 1 (25.0) | 3 (75.0) | 13.8 (1.7-280.8, p=0.02) | 8.3 (0.9-183.9, p=0.08) |
| HIV | Negative | 437 (81.4) | 100 (18.6) | - | - |
|  | Positive | 33 (86.8) | 5 (13.2) | 0.7 (0.2-1.6, p=0.40) | 0.6 (0.2-1.5, p=0.30) |
| Diabetes | No | 435 (81.9) | 96 (18.1) | - | - |
|  | Known | 67 (83.8) | 13 (16.2) | 0.9 (0.5-1.6, p=0.70) | 1.1 (0.5-2.3, p=0.80) |
| Smear<br>Microscopy | Negative | 82 (80.4) | 20 (19.6) | - | - |
|  | Positive | 421 (82.5) | 89 (17.5) | 0.9 (0.5-1.5, p=0.60) | 0.7 (0.4-1.4, p=0.30) |
| Smoking | Non-Smoker | 402 (82.7) | 84 (17.3) | - | - |
|  | Smoker | 99 (79.8) | 25 (20.2) | 1.2 (0.7-2.0, p=0.50) | 1.2 (0.7-2.3, p=0.50) |
| Alcohol Use | No | 355 (81.6) | 80 (18.4) | - | - |
|  | Yes | 148 (83.6) | 29 (16.4) | 0.9 (0.5-1.4, p=0.60) | 0.7 (0.4-1.2, p=0.20) |
| Lineage | 1 | 142 (87.7) | 20 (12.3) | - | - |
|  | 2 | 33 (73.3) | 12 (26.7) | **2.6 (1.1-5.8, p=0.02)** | 2.2 (0.9-5.3, p=0.07) |
|  | 3 | 236 (86.4) | 37 (13.6) | 1.1 (0.6-2.0, p=0.70) | 1.1 (0.6-2.0, p=0.80) |
|  | 4 | 92 (69.7) | 40 (30.3) | **3.1 (1.7-5.7, p<0.001)** | **3.3 (1.8-6.3, p<0.001)** |

In more detail at an SNS threshold ≤ 25 SNSs, we found nine clusters in L1 (7 pairs, 2 clusters of 3 isolates) spanning 20 isolates (12.3% of the 162 L1 isolates). For L2 we identified six transmission pairs of isolates (26.7% of the 45 L2 isolates). For L3, we identified 18 clusters (17 pairs and 1 cluster of 3 isolates) spanning 37 isolates (13.6% of the 273 L3 isolates). Lastly for L4, we identified 14 clusters (12 pairs and 2 networks containing 4 and 12 isolates) spanning 40 isolates (30.3% of the 132 L4 isolates).

We identified evidence supporting the transmission of drug resistance. Of the clustered isolates, 7.6% of L1, 50.0% of L2, 22.2% of L3 and 20% of L4 harbored resistance to one or more drugs (**Supplementary Table 5**). One L4 transmission network of four isolates (all ≤ 6 SNSs apart) tested levofloxacin mono-resistant and harbored the D94G mutation in *gyrA* (**Figure 4**). These four isolates with levofloxacin mono-resistance belonged to participants with no known epidemiological links. Two additional L4 pairs of isolates (≤10 SNSs apart) harbored isoniazid mono-resistance for a total of 20% of clustered isolates harboring drug resistance (n=40).

**Figure 4:**
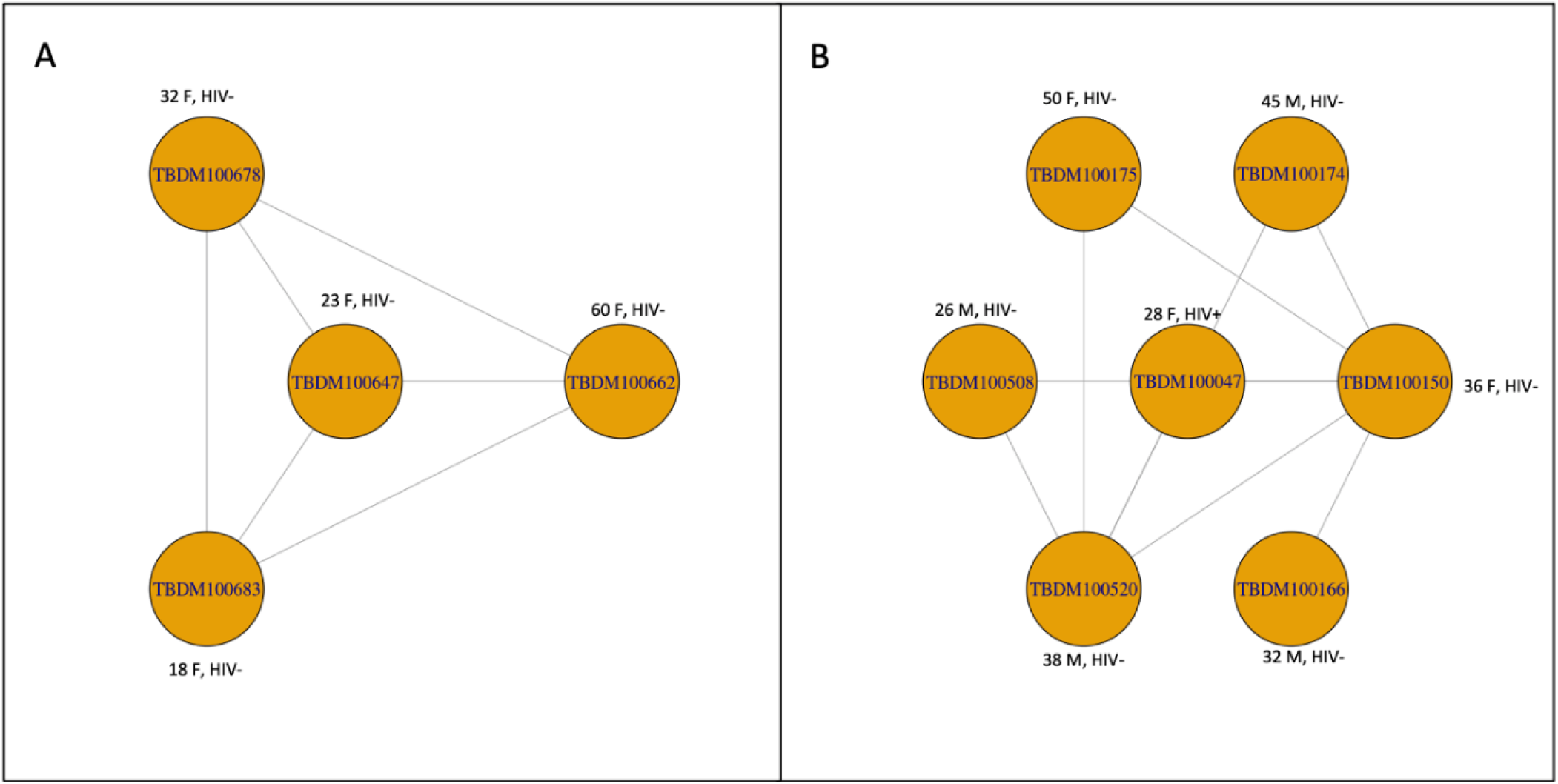
Largest network of genetically closely related lineage 4 isolates identified using a single nucleotide substitution (SNS) cut-off of ≤ 10 SNSs. Each vertex in this network is an isolate and each edge represents a SNS distance of ≤ 10. In (A) all isolates were rifampin-susceptible (genotypic and phenotypic) and levofloxacin-resistant based on genotypic prediction and were ≤ 5 SNSs apart except 678 and 662 that were 6 SNSs apart. In (B), all isolates were pan-susceptible based on genotype and phenotype. The numbers near each vertex depict age of study participant in years, gender (F: Female, M: Male) and human immunodeficiency virus co-infection status: positive (+) or negative (-).

### Frequency and Timing of Resistance Acquisition

L2 had a higher frequency of drug resistance acquisition (acquisition of new resistance-conferring mutations as % of total isolates) as compared to other lineages, specifically for rifampin, ethambutol, streptomycin, and levofloxacin (**Supplementary Table 6**). We investigated the specific drug-resistance conferring mutations acquired (**Table 4**) and the timing of these acquisitions with L2 demonstrating the highest number of resistance acquisition events.

**Table 4:** Acquisition of drug-resistance conferring mutations by lineage. Year of acquisition calculated based on median height of the branching node where resistance was acquired.

| Drug and mutation | Number of acquisitions and year(s) of acquisition |  |  |  |
| --- | --- | --- | --- | --- |
|  | Lineage 1 | Lineage 2 | Lineage 3 | Lineage 4 |
| **Rifampin** |  |  |  |  |
| *rpoB* S450L | - | 5 (2010, 2011, 2014 x2, 2016) | 3 (2000, 2003, 2011) | 2 (1996, 2015) |
| *rpoB* L430P | 1 (2010) | - | 1 (2002) | - |
| *rpoB* I491F | 1 (2012) | - | - | - |
| *rpoB* Q432K | - | - | 1 (2010) | - |
| **Isoniazid** |  |  |  |  |
| *katG* S315T | 6 (2007, 2008, 2009, 2011, 2012, 2014) | 6 (2009, 2014, 2015, 2016x2, 2017) | 15 (2000, 2002, 2003, 2005, 2006, 2007 x2, 2008, 2010, 2012, 2013, 2014, 2015, 2018 x2) | 5 (1996, 2011, 2014, 2015, 2018) |
| *fabG1-inhA* promoter C15T | 3 (1995, 2013, 2017) | 1 (2015) | 5 (2008, 2011, 2012, 2013x2) | 2 (2005, 2014) |
| *inhA* S94A | 1 (2010) | - | - | - |
| *katG* S315N | - | - | - | 2 (2007, 2014) |
| *fabG1-inhA* promoter G17T | - | - | - | 1 (2018) |
| **Ethambutol** |  |  |  |  |
| *embB* M306V | - | 5 (2009, 2014, 2016x3) | 2 (2010) | - |
| *embB* M306I | - | - | 1 (2002) | - |
| *embB* G406A | - | - | 1 (2009) | - |
| *embC* R738Q | - | - | 1 (2007) | - |
| *embA* E951D | - | 1 (2009) | - | - |
| *embB* Q497R | - | 1 (2016) | - | - |
| **Pyrazinamide** |  |  |  |  |
| *pncA* L27P | - | 1 (2016) | - | - |
| *pncA* i517C | - | - | 1 (2010) | 1 (2015) |
| *pncA* i391CC | - | - | 1 (2000) | - |
| *pncA* V139A | - | - | 1 (2008) | - |
| *pncA* P69S | - | - | 1 (2007) | - |
| *pncA* G97D | - | - | - | 1 (1999) |
| **Streptomycin** |  |  |  |  |
| *rrs* C517T | 1 (2012) | - | 1 (2013) | - |
| *rpsL* K88R | 1 (1995) | 2 (2014, 2015) | 1 (2014) | - |
| *gid* A138V and *rrs* T1208G | 1 (1995) | - | - | - |
| *gid* S70R | 1 (1994) | - | - | - |
| *rpsL* K43R | - | 6 (2014, 2015, 2016x2, 2017x2) | 2 (2000) | 1 (2007) |
| *gid* d351C | - | - | 4 (2004, 2010, 2014, 2018) | - |
| *gid* d87G | - | - | 1 (2005) | - |
| *gid* d115G | - | - | 1 (2012) | 1 (2013) |
| *rrs* A514C | - | - | 2 (2008, 2009) | 1 (2015) |
| *gid* P75S | - | - | - | 1 (2010) |
| *rrs* C936T, T979A, and A948T | - | - | - | 1 (2015) |
| *rrs* C936T, T958A, and A948T | - | - | - | 1 (2018) |
| *rrs* T958A, A948T, T327C and C1050T | - | - | - | 1 (1995) |
| *gid* L90R | - | - | - | 1 (2009) |
| *rpsL* K88M | - | 1 (2009) | - | - |
| *gid* E92D | - | 1 (2017) | - | - |
| **Levofloxacin** |  |  |  |  |
| *gyrA* A90V | 1 (1998) | 1 (2008) | 3 (2003, 2007, 2012) | 2 (2006, 2017) |
| *gyrB* M330I | 1 (2008) | - | - | - |
| *gyrA* D94G | - | 4 (2009, 2015, 2016, 2017) | 5 (2008, 2009, 2011x2, 2012) | 1 (2018) |
| *gyrA* D94N | - | 1 (2017) | - | - |
| *gyrA* D94A | - | 1 (2016) | 1 (2011) | - |
| *gyrA* D94Y | - | 1 (2016) | 1 (2009) | - |
| *gyrA* S91P | - | - | - | 2 (1924,<br>2016) |
| *gyrB* N538T | - | - | - | 1 (2018) |
| **Capreomycin** |  |  |  |  |
| *tlyA* H68R | - | - | 1 (2012) | - |

### Propensity to Propagate

We investigated host related factors associated with TB transmission for each host using a previously developed score called the Propensity to propagate (PTP)^44^ (**Methods**). The median PTP for all the 612 hosts was 1.02 (IQR: 0.88-1.17) and did not vary significantly by lineage (L1: median 1.01 [IQR: 0.88-1.17], L2: 1.17 [1.01-1.30], L3: 1.01 [0.88-1.17], L4: 1.06 [0.89-1.17], p > 0.1 for each pairwise test between lineages, **Figure 5**).

**Figure 5:**
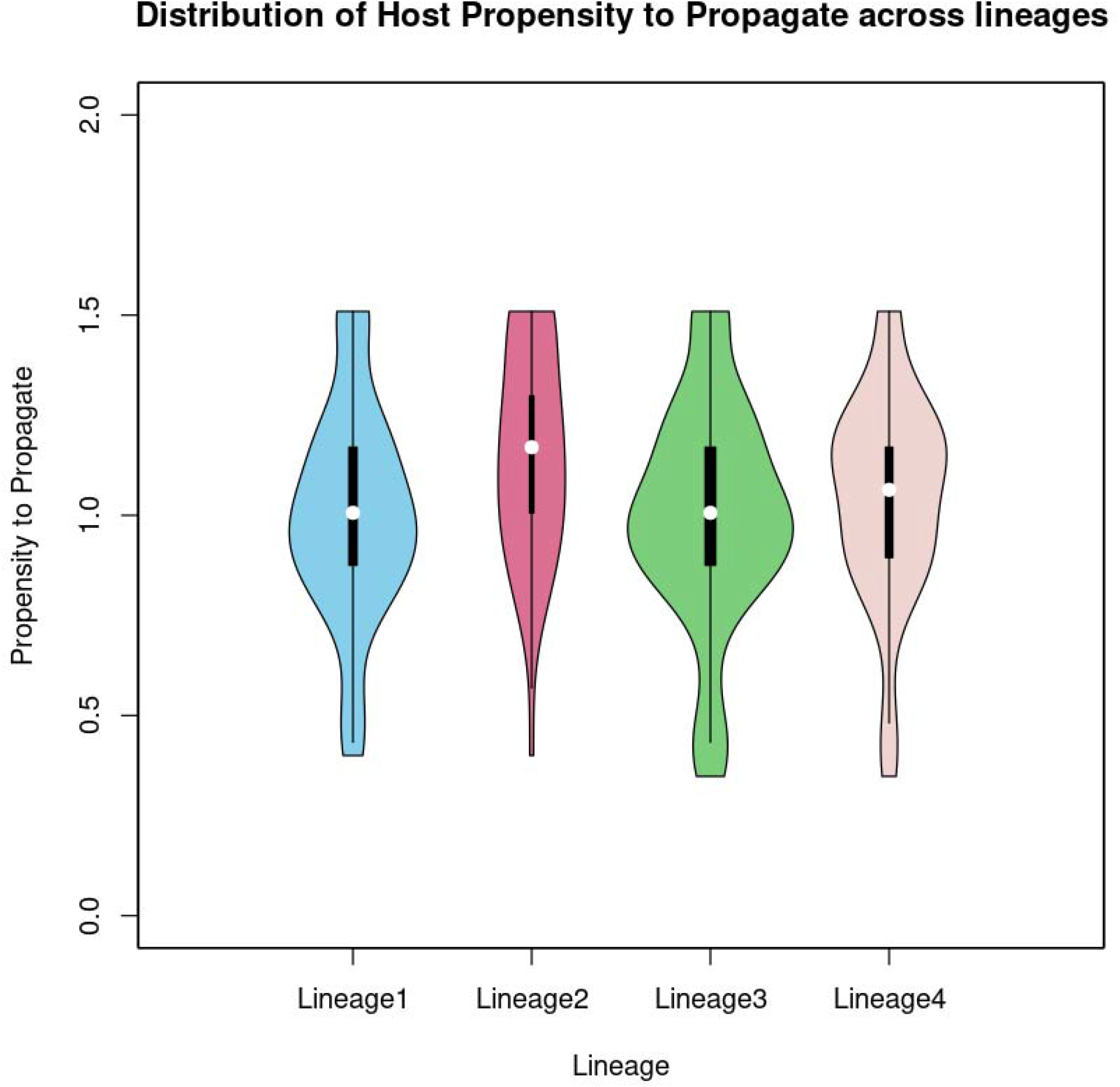
Lineage-wise distribution of host propensity to propagate (PTP). Subjects with isolates belonging to lineage 2 did not have a significantly higher PTP compared to other three lineages based on a one-sided two-sample Kolmogorov-Smirnov test

## Discussion

Using *Mtb* WGS of participants with PTB in western India, we characterize differences in transmission and resistance between the four major *Mtb* lineages. We find evidence of higher transmissibility of modern lineages L2 and L4 as compared to L1 and L3, consistent with findings from other parts of the world^8–10^. These differences persisted after controlling for host factors. Our results foreshadow the possible future displacement of native Indian lineages (L1 and L3) with modern lineages (L2 and L4) in western India unless TB control measures reduce transmission substantially. L2 was also noted to be more drug resistant, with more clustering and more resistance acquisition events, raising concern for its expansion fueling the problem of drug resistant TB in India. We identify a high rate of discordance of genotype and phenotype-based resistance diagnosis for TB isolates from Pune. Genotypic prediction increased the sensitivity of resistance diagnosis, with all resistant missed by phenotype harboring known/canonical resistance variants. Yet, a substantial proportion of isolates predicted susceptible were measured to be phenotypically resistant; among these we identified several possible resistance-candidate variants not otherwise documented in the WHO resistance catalogue. Identification of novel resistance-associated mutations may put available genotype-based assays for resistance detection at risk, accentuating the critical need for additional studies in western India.

Among the anti-TB drugs, fluoroquinolones (FQs) deserve special attention. Since multidrug resistant (MDR) TB was first recognized, FQs have been used for its treatment^47^. Recent clinical trials also support their use in treating drug susceptible (DS) TB to shorten the treatment duration from six to four months^48,49^. The bactericidal activity of FQs towards MTB is through targeting of deoxyribonucleic acid (DNA) gyrase, an enzyme necessary for DNA replication, encoded by gyrase genes^50^. FQ-R in *Mtb* is acquired through mutations in these essential gyrase genes. Hence, as has been seen in other bacteria^51^, it is suspected, but not studied clinically, that these mutations would confer a fitness cost on *Mtb*, lowering transmissibility. But the rising rates of resistance to FQs (FQ-R) in several countries^1^, including India^13,52–54^, are concerning for acquisition of compensatory mutations, that allow these strains to overcome the fitness cost, hypothetically even conferring an increased transmissibility advantage^55^. In our study, we identified a group of closely related rifampin susceptible isolates that harbored the D94G mutation in the *gyrA* gene, known to confer FQ-R^45^ and is consistent with independent recent work reporting evidence for transmission of ‘fit’ FQ-R isolates in other parts of the world. Over time, this may result in an increase in FQ-R rates^56^ and supports the need for heightened surveillance, including comprehensive DST for regimen selection^57^.

This study had several limitations. We used a convenience sample of participants seeking care that can introduce bias, miss transmission links and lead to incomplete sampling of certain lineages. However, lineage assignments were unknown at the time of subject enrolment, so sampling is unlikely to affect our findings. The use of WGS does not always allow inference of direction of transmission and misses genetic diversity in approximately 10% of the genome^58,59^ underestimating the genetic distance between isolates but this should impact all isolates equally, supporting our differential clustering finding between lineages.

In conclusion, our findings highlight that there are inherent differences between *Mtb* lineages with implications for TB control, surveillance, and monitoring. As modern and more drug resistant lineages take further hold in India, the proportion of TB with drug resistance may continue to rise, along with the number of possible new resistance associated variants. To achieve control, resources will need to be directed towards interrupting transmission by increasing efforts towards active case finding, contact tracing, early diagnosis and treatment. The wider adoption of WGS can assist these efforts by providing quicker and more comprehensive genotype-based DST results allowing clinicians to tailor therapy sooner and in turn decreasing transmission.

## Supporting information

Supplementary Results

## Data Availability

All data produced in the present study are available upon reasonable request to the authors

## Acknowledgements

The authors wish to thank the study participants and acknowledge the clinical staff who collected and maintained samples, and the laboratory staff who conducted the culturing, phenotyping and genotyping work across multiple institutions.

## Author’s contributions

The study question and analysis design was conceived by AD and MF. Analysis was executed by AD with key contributions from YE, LF, MG. AD and MF had access to the data and are responsible for study integrity. VM led the field participant recruitment, data collection, sequencing coordination, and secured grant funding with contributions from AG, JG, AK, RK, RL, NG, NP, JAT, MP, SD and DK. DME and MS performed the sequencing. AD and MF wrote the manuscript. All authors reviewed the manuscript and provided edits.

