## Supplementary Results for "Modern lineages of *Mycobacterium tuberculosis* were recently introduced in western India and demonstrate increased transmissibility"

Avika Dixit^1,2^, Anju Kagal^3^, Yasha Ektefaie^2^, Luca Freschi^2^, Rajesh Karyakarte^3^, Rahul Lokhande^3^, Matthias Groschel^2^, Jeffrey A Tornheim^4,5^, Nikhil Gupte^4,7,8^, Neeta N Pradhan^7,8^, Mandar S Paradkar^7,8^, Sona Deshmukh^7,8^, Dileep Kadam^3^, Marco Schito^9^, David M. Engelthaler^10^, Amita Gupta^4,6^, Jonathan Golub^5^, Vidya Mave*^4,7,8^, Maha Farhat*^2,11^

*These authors contributed equally to this work

^1^Division of Infectious Diseases, Boston Children's Hospital, Boston MA, USA

^2^Department of Biomedical Informatics, Harvard Medical School, Boston MA, USA

^3^Byramjee-Jeejeebhoy Government Medical College, Pune, India

^4^Center for Clinical Global Health Education, Division of Infectious Diseases, Department of Medicine, Johns Hopkins University School of Medicine, Baltimore, MD, USA

^5^Center for Tuberculosis Research, Department of Medicine, Johns Hopkins University School of Medicine, Baltimore, MD, USA

^6^Department of International Health, Johns Hopkins Bloomberg School of Public Heath, Baltimore, MD, USA

^7^Byramjee-Jeejeebhoy Medical College-Johns Hopkins University Clinical Research Site, Pune, India

^8^Johns Hopkins India, Pune, India

^9^Critical Path Institute, Tucson, AZ, USA

^10^Translational Genomics Research Institute, Flagstaff, AZ, USA

^11^Division of Pulmonary and Critical Care Medicine, Massachusetts General Hospital, Boston, MA, USA

**Supplementary Table 1:** Phenotypic drug susceptibility testing results.

| Drug | N tested | Resistant n (%) |
| --- | --- | --- |
| Isoniazid | 574 | 73 (12.7) |
| Rifampin | 574 | 30 (5.2) |
| Ethambutol | 573 | 29 (5.06) |
| Streptomycin | 573 | 61 (10.7) |
| Multidrug resistant (MDR, i.e., resistant to both isoniazid and rifampin) | 574 | 24 (4.2) |

**Supplementary Table 2:** **Mutations identified in isolates predicted to be genotypically resistant but that tested susceptible on phenotypic drug susceptibility testing results.** Isolates were not tested for phenotypic resistance to second line agents. Only the four isolates predicted as capreomycin-resistant by genotype were tested for phenotypic resistance to capreomycin.

| Drug | Discrepant (Geno-R Pheno#1-S) Isolates (n) | Mutations (Geno-R Pheno#1-S) | Discrepant (Geno-R Pheno#2-S) Isolates (n) | Mutations (Geno-R Pheno#2-S) |
| --- | --- | --- | --- | --- |
| Isoniazid | 19^a^ | S315T katG (n = 13)  S315N katG (n = 1)  C15T fabG1-inhA promoter (n = 5) | 3^a^ | S315T katG (n = 3) |
| Rifampin | 5^b^ | S450L rpoB (n = 3)  L430P rpoB (n = 2) | 1^b^ | L430P rpoB (n = 1) |
| Ethambutol | 6^c^ | M306V embB (n = 4, of which 2 also had T1082A embB and 1 had R738Q embC)  M306I embB (n = 1)  R738Q embC (n = 1)  E951D embA (n = 1) | 3^c^ | M306V embB (n = 1, the isolate also harbored R738Q embC)  M306I embB (n = 1, the isolate also harbored R738Q embC)  E951D embA (n = 1) |
| Streptomycin | 13^d^ | K88R rpsL (n = 1)  K43R rpsL & E92D gid (n = 4)  d351C gid (n = 3)  C936T rrs & A948T rrs (n = 2, of which one also had T958A rrs and another also had T979A rrs)  L90R gid (n=1)  T1208G rrs & A138V gid (n=1)  T327C rrs, A948T rrs, T958A rrs & C1050T rrs (n=1) | 4^d^ | d351C gid (n = 2)  C936T rrs & A948T rrs (n = 2, of which one also had T958A rrs and another also had T979A rrs) |
| Capreomycin | 4 | H68R tlyA | - | - |

**Geno-R Pheno#1-S:** Isolates that were determined to be resistant using random forest prediction (genotypic) but tested susceptible on initial phenotypic drug susceptibility testing (DST)

**Geno-R Pheno#2-S:** Isolates that were determined to be resistant using random forest prediction (genotypic) but tested susceptible on both initial and repeat phenotypic DST

^a^Only 10 of the 19 isolates were available for repeat DST

^b^Only 1 of the 5 isolates was available for repeat phenotypic DST

^c^Only 4 of the 6 isolates were available for repeat phenotypic DST

^d^Only 7 of the 13 isolates were available for repeat phenotypic DST

**Supplementary Table 3: Mutations identified in isolates predicted to be genotypically susceptible but that tested resistant on phenotypic drug susceptibility testing results.** None of these mutations have been associated with drug resistance based on the recently released WHO resistance catalogue (https://www.who.int/publications/i/item/9789240028173). Only non-synonymous mutations are listed.

| Drug | Mutation | Discrepant (Geno-S Pheno#1-R) Isolates with mutation (n) | Discrepant (Geno-S Pheno#2-R) Isolates with mutation (n) | Association with Resistance (based on WHO catalogue 2021) |
| --- | --- | --- | --- | --- |
| Isoniazid  Geno-S Pheno#1-R Isolates (n=35)  Geno-S Pheno#2-R Isolates (n=3)^a^ | SNP_CN_2154724_C1388A_R463L_katG | 30 | 2 | Not Associated |
|  | SNP_P_2726105_G88A_promoter.ahpC | 17 | 2 | Not Associated |
|  | SNP_CN_471666_A974G_M325T_ndhA | 11 | - | Not found |
|  | SNP_P_409297_A65G_promoter.iniB.iniA.iniC | 3 | - | Not found |
|  | DEL_I_472712_d69TTGTGGGCC_inter.ndhA.Rv0393 | 3 | - | Not found |
|  | SNP_CN_471640_C1000T_A334T_ndhA | 2 | - | Not found |
|  | SNP_CN_2102240_C803T_R268H_ndh | 2 | - | Not Associated |
|  | DEL_P_2726159_d34A_promoter.ahpC | 1 | - | Not found |
|  | INS_I_472712_i69TTGTGGGCC_inter.ndhA.Rv0393 | 1 | - | Not found |
|  | SNP_CN_1674956_C755T_A252V_inhA | 1 | - | Not found |
|  | SNP_CN_2518894_G780C_K260N_kasA | 1 | - | Not found |
|  | SNP_CN_2519353_C1239A_F413L_kasA | 1 | - | Not found |
|  | SNP_CN_2726418_G226A_E76K_ahpC | 1 | - | Not found |
|  | SNP_CN_410104_T743C_V248A_iniB | 1 | 1 | Not found |
|  | SNP_CN_410191_C830T_A277V_iniB | 1 | - | Not found |
|  | SNP_CN_411298_A461C_E154A_iniA | 1 | - | Not found |
|  | SNP_CN_412071_G1234A_E412K_iniA | 1 | - | Not found |
|  | SNP_CN_4247136_C623A_T208N_embB | 1 | - | Not found |
|  | SNP_CN_4247368_C855A_F285L_embB | 1 | - | Not found |
|  | SNP_CN_4247680_G1167A_M389I_embB | 1 | - | Not found |
|  | SNP_CN_4248225_C1712T_A571V_embB | 1 | - | Not found |
|  | SNP_CN_4248755_G2242A_G748R_embB | 1 | - | Not found |
|  | SNP_CN_4249019_G2506A_G836R_embB | 1 | - | Not found |
|  | SNP_CZ_411504_C667T_Q223._iniA | 1 | - | Not found |
|  | SNP_CZ_471616_G1024A_R342._ndhA | 1 | 1 | Not found |
|  | SNP_P_1673432_T8A_promoter.fabG1.inhA | 1 | - | Uncertain Significance |
| Rifampicin  Geno-S Pheno#1-R Isolates (n=15)  Geno-S Pheno#2-R Isolates (n=0)^b^ | SNP_CN_762523_C2717A_P906Q_rpoB | 1 | - | Not found |
|  | SNP_P_759546_A261G_promoter.rpoB | 1 | - | Not Associated |
|  | SNP_P_759642_C165T_promoter.rpoB | 1 | - | Not found |
|  | SNP_P_759746_C61T_promoter.rpoB | 5 | - | Not Associated |
| Ethambutol  Geno-S Pheno#1-R Isolates (n=20)  Geno-S Pheno#2-R Isolates (n=1)^c^ | SNP_CN_1417019_C329T_C110Y_embR | 6 | 1 | Not Associated |
|  | SNP_I_1417554_G104C_inter.embR.Rv1268c | 6 | 1 | Not found |
|  | SNP_P_409297_A65G_promoter.iniB.iniA.iniC | 2 | - | Not found |
|  | SNP_CN_412071_G1234A_E412K_iniA | 1 | - | Not found |
|  | SNP_CN_4243948_G716A_R239Q_embA | 1 | - | Not found |
| Streptomycin  Geno-S Pheno#1-R Isolates (n=33)  Geno-S Pheno#2-R Isolates (n=4)^d^ | SNP_I_1471659_C187T_inter.murA.rrs | 33 | 4 | Not Associated |
|  | SNP_P_781395_T165C_promoter.rpsL | 32 | 4 | Not Associated |
|  | SNP_CN_4407848_C355T_A119T_gid | 2 |  | Not found |
|  | SNP_CZ_4407686_C517A_E173._gid | 2 | 1 | Not found |
|  | DEL_CF_4408101_d102C_35_gid | 1 | 1 | Not found, *gid* Del nt103 associated with resistance |
|  | SNP_CN_4407604_G599A_A200V_gid | 1 | - | Not found |
|  | SNP_CN_4407866_C337G_E113Q_gid | 1 | 1 | Not found |
|  | SNP_CN_4407917_G286A_R96C_gid | 1 | - | Not found |
|  | SNP_CN_4407922_A281G_L94P_gid | 1 | - | Not found |
|  | SNP_CN_4407929_C274G_E92Q_gid | 1 | 1 | Not found, *gid* E92D not associated with resistance |
|  | SNP_CN_4408051_T152G_N51T_gid | 1 | 1 | Not found |
|  | SNP_CN_4408144_C59G_R20P_gid | 1 | - | Not found |
|  | SNP_CZ_4408025_C178A_E60._gid | 1 | - | Not found |
|  | SNP_P_4408266_G68T_promoter.gid | 1 | - | Not found |

**Geno-S Pheno#1-R:** Isolates that were determined to be susceptible using random forest prediction (genotypic) but tested resistant on initial phenotypic drug susceptibility testing (DST)

**Geno-S Pheno#2-R:** Isolates that were determined to be susceptible using random forest prediction (genotypic) but tested resistant on both initial and repeat phenotypic DST

^a^Only 23 of the 35 isolates were available for repeat DST

^b^Only 11 of the 15 isolates was available for repeat phenotypic DST

^c^Only 13 of the 20 isolates were available for repeat phenotypic DST

^d^Only 25 of the 33 isolates were available for repeat phenotypic DST

**Supplementary Table 4: Number of genetically similar isolates by lineage based on SNP cut-offs**

| SNPs  Lineage | **<= 5** | **<= 10** | **<= 15** | **<=25** |
| --- | --- | --- | --- | --- |
| **1**  (n=162) | 2 pairs  (2.5%) | 4 pairs  (4.9%) | 4 pairs  1 network (3 isolates)  (6.8%) | 7 pairs  2 networks (3 isolates each)  (12.3%) |
| **2**  (n=45) | 1 pair  (4.4%) | 1 pair  (4.4%) | 3 pairs  (13.3%) | 6 pairs  (26.7%) |
| **3**  (n=273) | 1 pair  (0.7%) | 9 pairs  (6.6%) | 11 pairs  (8%) | 17 pairs  1 network (3 isolates)  (13.5%) |
| **4**  (n=132) | 7 pairs  2 networks (4 & 6 isolates)  (18.2%) | 8 pairs  2 networks (4 & 7 isolates)  (20.5%) | 10 pairs  2 networks (4 & 7 isolates)  (23.5%) | 12 pairs  2 networks (4 & 12 isolates)  (30.3%) |

**Supplementary Table 5: Resistant and clustered isolates (separated by 25 or fewer SNSs)**

| **Lineage** (n clustered, see Table 1 for total sample size by lineage) | **Number of clustered isolates with resistance** (≤25 SNPs apart) | **Resistance pattern** |
| --- | --- | --- |
| L1 (n=26) | 2 (7.6%) | - one pair with isoniazid and ethionamide resistance |
| L2 (n=12) | 6 (50.0%) | - one pair with isoniazid mono-resistance - one pair with MDR (and also resistant to ethambutol, streptomycin and fluoroquinolones) - one pair with streptomycin and fluoroquinolone resistance but susceptible to isoniazid and rifampin |
| L3 (n=18) | 4 (22.2%) | - both pairs were isoniazid mono-resistant - one pair also harboring streptomycin resistance   (both pairs were ≤10 SNPs apart) |
| L4 (n=40) | 8 (20.0%) | - One networks (4 isolates, all ≤ 6 SNPs apart) were levofloxacin mono-resistant harboring the D94G mutation in *gyrA*. - Two pairs of isolates (≤10 SNPs apart) with isoniazid mono-resistance |

**Supplementary Table 6:** **Phylogenetic branches with drug resistance acquisition across each of the four major lineages**. Sample sizes (N) represent the number of tips/isolates in each phylogeny.

|  | Lineage 1  (N = 162)  n (%) | Lineage 2  (N = 45)  n (%) | Lineage 3  (N 273)  n (%) | Lineage 4  (N 132)  n (%) |
| --- | --- | --- | --- | --- |
| Rifampin | 2 (1.2) | 5 (11.4) | 5 (1.8) | 2 (1.6) |
| Isoniazid | 10 (6.2) | 7 (15.9) | 20 (7.4) | 10 (7.7) |
| Ethambutol | 0 (0) | 7 (15.9) | 5 (1.8) | 0 (0) |
| Pyrazinamide | 0 (0) | 1 (2.2) | 4 (1.5) | 2 (1.6) |
| Streptomycin | 4 (2.5) | 10 (22.7) | 12 (4.4) | 8 (6.1) |
| Capreomycin | 0 (0) | 0 (0) | 1 (0.4) | 0 (0) |
| Levofloxacin | 2 (1.2) | 8 (18.2) | 10 (3.7) | 6 (4.6) |
